# Fungal pleural infection due to *Microascus gracilis* with pulmonary aspergillosis after COVID-19 pneumonia

**DOI:** 10.1101/2023.12.07.23298951

**Authors:** Zhi-Min Hu, Li-Na Mao, Ti-Ying Deng, Bin-Tao Su, Yi Yang, Bi-Lin Dong, Qing Xu, Shuo Yang

## Abstract

**Background:** *Scopulariopsis/Microascus* is a rare but devastating pathogen due to its intrinsic resistance to nearly all available antifungal agents. *Microascus gracilis*, an ascomycetous mould in the order *Microascales*, family *Microascaceae*, has recently emerged as a significant invasive pathogen causing opportunistic infections.

**Objectives and Methods:** We present a case of pleural infection caused by *M. gracilis* with pulmonary aspergillosis in an immunocompromised man after COVID-19 pneumonia. To further understand the characteristics of the pathogen isolated from the patient, we identified the strain through mycological characteristics, matrix-assisted laser desorption/ionization (MALDI) time-of-flight (TOF) mass spectrometry (MALDI-TOF MS) and internal transcribed spacer (ITS)-based sequencing, and performed *in vitro* drug susceptibility testing against common antifungal agents. Moreover, we assessed lymphocyte subsets and programmed cell death protein 1 (PD-1) expression in peripheral blood and pleural effusion to monitor the efficacy of therapy with thymosin-α-1 and intravenous immunoglobulin.

**Results:** Filamentous fungi isolated from pleural fluid were identified as *M. gracilis* based on classical morphology, mass spectrometry and molecular biology methods. The susceptibility results *in vitro* revealed that multiple antifungal agents were inactive against the strain. Adjuvant immunomodulatory treatment successfully increased the levels of CD3+ T and CD4+ T cells while decreasing the levels of CD3+PD-1+ and CD4+PD-1+ T cells in both peripheral blood and pleural effusion.

**Conclusions:** The immunocompromised host with opportunistic *M. gracilis* infection, rapid and accurate recognition through direct microscopic testing with calcofluor white and MOLDI-TOF MS, is the key to achieving a definite diagnosis, and a combination of antifungal therapy with immunomodulatory therapy is vital for improving survival.

## 1 INTRODUCTION

Pleural infection is a millennium-spanning condition that has been shown to be challenging to treat over many years.^1^ Fungi are uncommon causes of pleural infection and represent 3% of isolated pathogens,^2^ but there should be a strong suspicion of underlying dormant fungal infection for differential diagnosis when episodes of pleural effusions are encountered. Fungal infections tend to occur in patients with underlying immunocompromising conditions, and the most common isolated are *Candida* spp.^3^ As already well documented in medical articles, both immunocompromised and immunocompetent hosts are at an increased risk for developing fungal infections, most commonly *Candida* spp. and *Aspergillus* spp., but other opportunistic filamentous fungi have been emerging in recent years.^4^ In particular, even though infections of the genera *Scopulariopsis*/*Microascus* are very rare, as already reviewed, they are associated with high morbidity and mortality in the clinical setting due to delayed diagnosis and intrinsic resistance to currently available antifungal agents.^4^ These fungi are commonly isolated from soil, air, decaying organic material, dung, insects and moist indoor environments.^4–10^ *M. gracilis*, the species of the genus *Microascus*, was first isolated in 1962 from food in Japan by Inagaki.^11^ In the past, it was known as *Scopulariopsis gracilis*, which has been recategorized and designated *M. gracilis*.^10^ Due to the increase in predisposing factors in parallel with medical technological advances, the number of cases caused by these organisms has been on the rise.

## 2 **CASE REPORT**

An old male presented to the hospital with cough and yellow phlegm lasting for one week followed by fever and progressive dyspnoea that developed 2 days prior to admission. In December 2022, coronavirus disease 2019 (COVID-19) was diagnosed by RT‒PCR for severe acute respiratory syndrome coronavirus (SARS-CoV-2) and recovered with therapeutic strategies for COVID-19 pneumonia. Two month after discharge, the patient was diagnosed with nephrotic syndrome and received treatment with an immunosuppressive regimen of prednisone and cyclophosphamide. The patient reported a history of hypertension for almost 10 years, and his blood pressure was controlled well by using amlodipine and metoprolol. The patient’s underlying conditions included chronic obstructive pulmonary disease (COPD), interstitial pneumonia, and steroid-induced diabetes. On physical examination, he was revealed to be sick with wheezing and subcostal retraction requiring supplemental oxygen delivered by standard nasal cannula. There was no clinical evidence of onychomycosis or paronychia. At presentation, his body temperature was 37.6 °C, his blood pressure was 142/88 mmHg, and his pulse was 92 bpm. His blood work showed an elevated leukocyte count of 13.43×10^9^ cells/L, haemoglobin 137 g/L, platelets 131×10^9^/L, high sensitivity c-reactive protein (hsCRP) 11.1 mg/L, erythrocyte sedimentation rate (ESR) 19 mmHg, (1,3)-beta-d-glucan (G test) 100.65 pg/mL, and IL-6 11.94 pg/mL. Serologies for the galactomannan antigen test (GM test), syphilis, acquired immunodeficiency syndrome, and hepatitis A–E were negative. Blood and urine cultures were sterile. Chest computed tomography (CT) examination showed a new right-sided pneumothorax, and ground-glass opacities (GGOs) were diffusely distributed in both lungs, especially in the middle and lower lobes of the right lung (Figure 1A).

**FIGURE 1.**
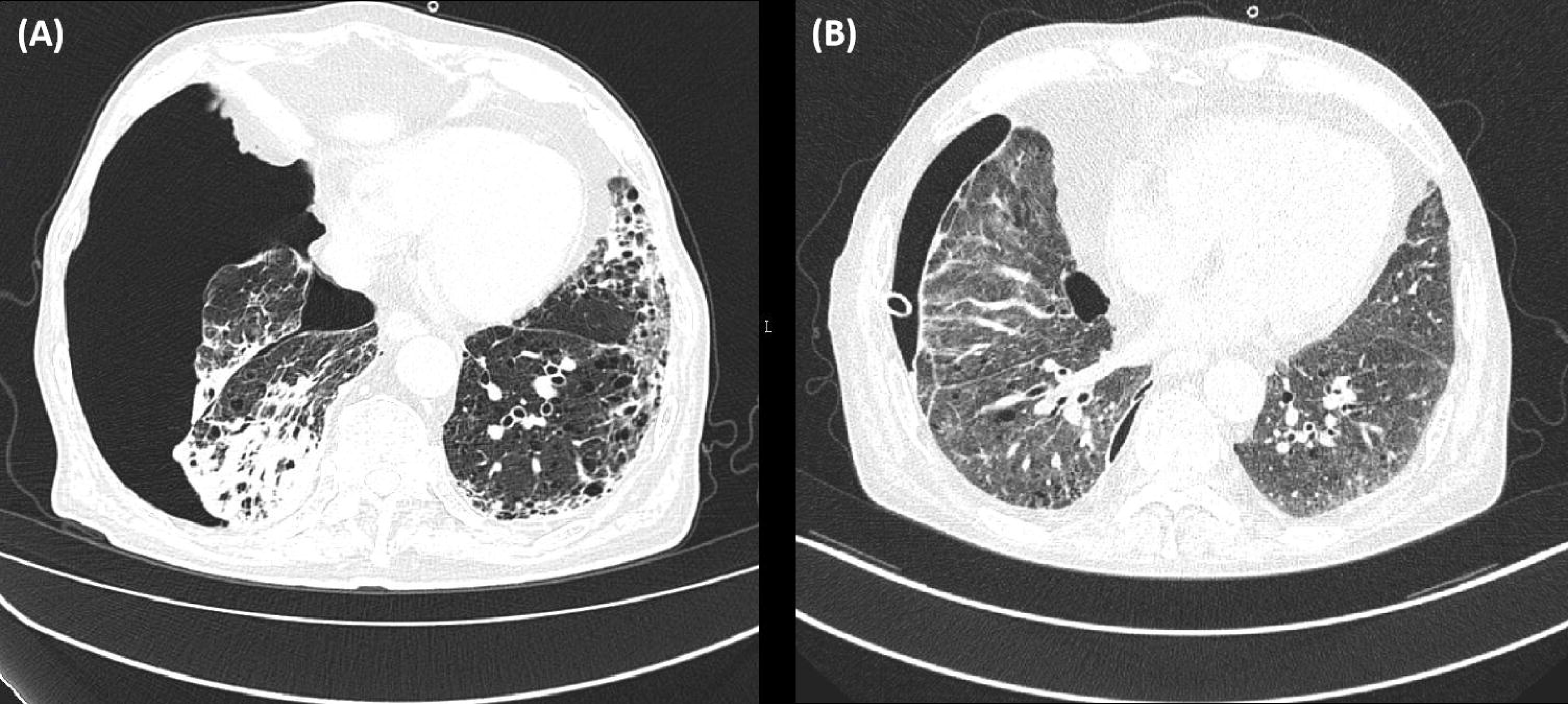
Chest computed tomography showed a new pneumothorax on the right lung, and ground-glass opacities were diffusely distributed in both lungs (A); a drainage tube, a cavity with an air-crescent sign and reduced hydropneumothorax in the right pleural cavity (B)

The timeline of diagnosis and targeted therapy is shown in Table 1. On Day 1, thoracentesis was performed, and the chest tube drainage system was maintained in an upright position below the patient’s chest to facilitate drainage. After that, the patient had recurrent infections in the lungs, and *Acinetobacter baumannii* and *Aspergillus fumigatus* were detected in sputum. Pulmonary symptoms were usually reduced or alleviated by empirical broad-spectrum antibiotics, including ceftriaxone/tazobactam, levofloxacin, fluconazole (FLZ) and voriconazole (VRC), for approximately 3 weeks. On Day 32, *Enterococcus faecalis* was incubated in the pleural fluid, and anti-biotherapy was adjusted to vancomycin for 3 weeks. The signs and symptoms of spontaneous hydropneumothorax were improved. On Day 65, the pathogen causing pleural reinfection was *Streptococcus sanguini*s, and the therapeutic agent was switched to ceftriaxone/tazobactam for 2 weeks. On Day 73, the symptoms of pneumonia and empyema were alleviated, and he was discharged home with intercostal drainage (ICD) *in situ*. Eight days later, he was readmitted because of worsening of dyspnoea, cough and wheezing, and no fever. According to the patient’s recollection, the chest drain was unsecure for fixation and accidental removal in house. His blood work showed a low absolute lymphocyte count 320/L. Abnormal laboratory findings included elevated hsCRP 189.3 mg/L and peripheral blood cytomegalovirus 1070 copies/ml. Repeated CT revealed a drainage tube, a cavity with an air-crescent sign and reduced hydropneumothorax in the right pleural cavity (Figure 1B). *Klebsiella pneumoniae* was detected in sputum, and anti-biotherapy was switched to latamoxef and valganciclovir for two weeks. After unplanned extubation, the new pleural catheter was reintubated away from the original region (Figures 2A). The culture from the removed chest tube was positive for mould growth. From Day 89 to Day 93, multiple cultures of pleural fluid from the new tube were positive for the same mould. Calcofluor white staining revealed many pigmented, septate fungal hyphae (Figures 2B) of the pleural fluid and septate fungal hyphae of BALF. In addition, the sample was separately inoculated onto Sabouraud’s dextrose agar (SDA) at 35 °C. The T-spot test and MTB/RIF GeneXpert detection assays were negative. Both G and GM tests were negative in serum and pleural fluid. However, the antifungal strategy of caspofungin (CAS, 50 mg/day) and posaconazole (POS, 0.3 g/day) was initiated despite the discouraging result of *in vitro* susceptibilities, and improvement of pleural effusion was achieved after 5 weeks of treatment. No adverse events were reported. On Day 95, intermittent fever was observed after the administration of the combined antifungal therapy. The serum and pleural fluid GM tests were still negative, and blood cultures were all sterile. A quantitative real-time PCR assay was developed to measure the human cytomegalovirus (HCMV) DNA load in peripheral blood, which was 2589 copies/ml. The BALF collected for metagenomics next generation sequencing (mNGS) showed high sequence numbers and relative abundance of *Pneumocystis jirovecii*, *Aspergillus fumigatus*, HCMV and EBV. The BALF GM antigen test was positive. With regard to the patient’s immunological and infectious condition, clindamycin 1.2 g/day, thymosin-α-1 4.8 mg/week and intravenous immunoglobulin 400 mg/kg/week were added. On Day 119, the symptoms of the lungs and pleural infection were relieved, the chest tube was removed, and the culture of the pleural catheter, pleural effusion and sputum were all negative. On Day 146, the nucleic acid of cytomegalovirus was normal, and the high level of CRP and persistent low level of absolute lymphocyte count were relieved. A chest CT scan showed that bilateral lung infection and pneumothorax were partly absorbed, lung tissue compression was less than 3%, and pleural effusion was reduced. The patient was discharged with no chief complaints and switched to terbinafine (TEB) 250 mg/day combined with POS 0.3 g/day for 4 weeks. After 3 months of follow-up, the patient had no obvious discomfort.

**FIGURE 2.**
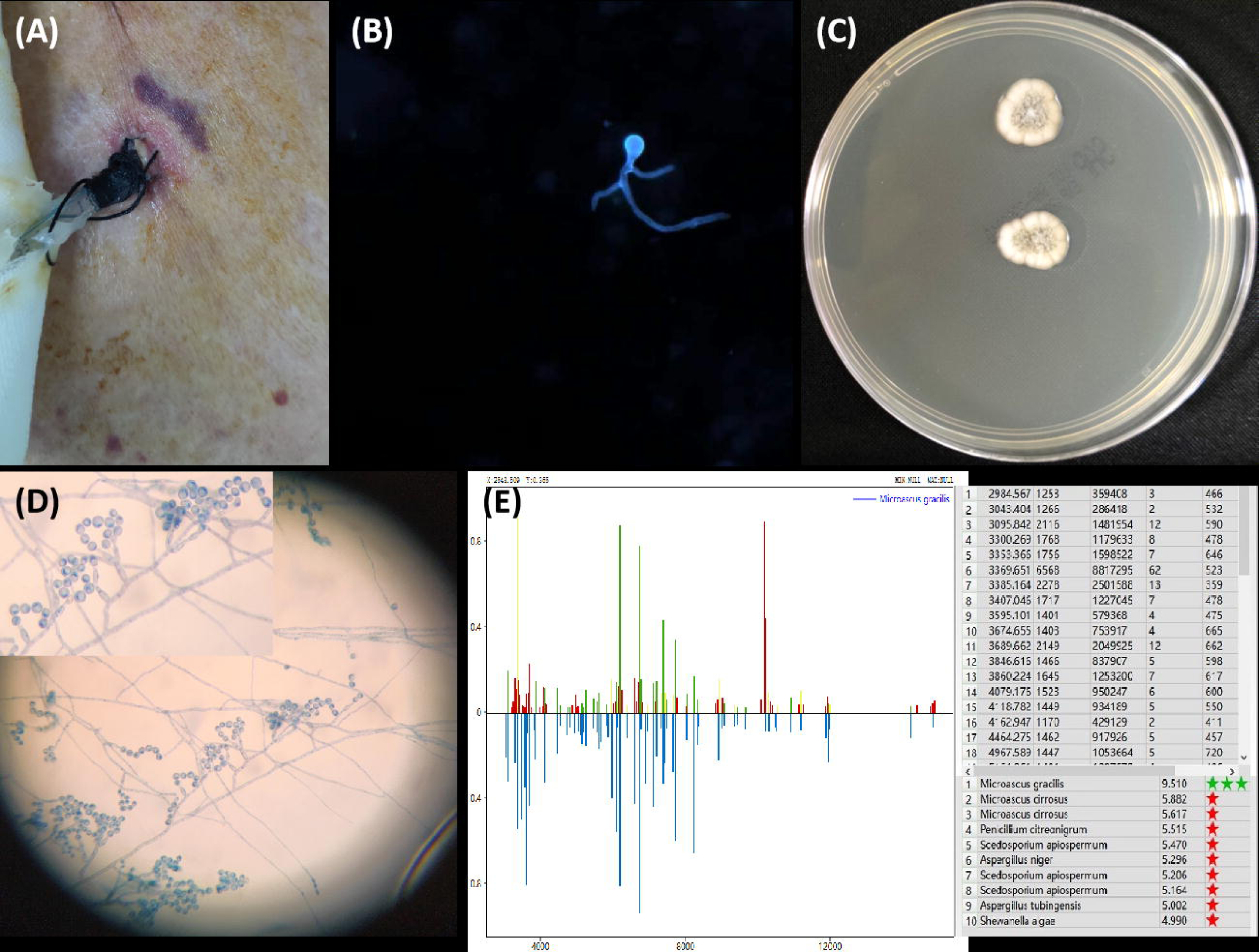
The image showed the new intercostal catheter on the right-sided chest(A). Calcofluor white staining showed the pigmented, septate fungal hyphae of pleural effusion, ×400 (B). The picture displays *Microascus gracilis* on Sabouraud’s dextrose agar after 5 days of incubation at 35 °C (C). Microscopic examination showing hyaline septate hyphae with bottle-shaped conidiogenous cells along with obovate-shaped, smooth conidia arranged in short chains, lactophenol cotton-blue staining, ×400 (D). The characteristic spectrometry of *M. gracilis* identified by MOLDI-TOF MS (E).

**TABLE 1.** Timeline of events.

| Time | Clinical features | Biology results | Therapy strategies |
| --- | --- | --- | --- |
| D1 | Progressive dyspnoea | Neutrophilia, abnormal chest CT scan | Chest tube insertion |
| D6 | Fever | *Acinetobacter baumannii* detected in sputum, High level of CRP, SAA, IL-6 and (1,3)-beta-d-glucan, low absolute lymphocyte count | Cefoperazone-sulbactam 9 g/day IV and prophylactic therapy fluconazole 0.2 g/day p.o. |
| D10 | Pneumonia and cough | *Aspergillus fumigate* detected twice in sputum | Levofloxacin 0.4 g/day and cefoperazone-sulbactam 9 g/day IV combined antifungal therapy (voriconazole 0.4 g/day p.o.) |
| D32 | Pleurisy | *Enterococcus faecalis* detected in pleural fluid, High level of CRP and IL-6, low absolute lymphocyte count | Switched to vancomycin 2 g/day IV |
| D65 | Increased pleural effusions | *Streptococcus sanguinis* detected in pleural fluid | Adjusted to cefoperazone-sulbactam 9 g/day IV |
| D73 | Outpatient | Negative sputum and pleural fluid |  |
| D81 | Readmitted due to worsening of the dyspnoea | High level of CRP and glycaemia, abnormal chest CT scan |  |
| D87 | Extubation and indwelling a new pleural catheter, cough | Cytomegaloviremia and persistent low level of absolute lymphocyte count; *Klebsiella pneumoniae* detected in sputum | Switched to latamoxef 4 g/day and valganciclovir 1.8 g/day p.o. |
| D89 | Pleural effusion appeared grossly turbid and yellowish | Filamentous fungi detected in pleural fluid, *Microascus gracilis* cultured in chest tube and pleural fluid | Switched antifungal therapy (caspofungin 50 mg/day IV and posaconazole 0.3 g/day p.o.) |
| D99 | Relieve respiratory symptoms | BALF mNGS detected *Pneumocystis jirovecii*, *Aspergillus fumigate*, HCMV and EBV; peripheral blood HCMV DNA positive; BALF GM test positive | Adding clindamycin 1.2 g/day IV, thymosin-α-1 4.8 mg/week and intravenous immunoglobulin 400 mg/kg/week |
| D119 | Remove chest tube | Chest tube, pleural fluid and sputum culture were negative | Switched antifungal therapy (terbinafine 250 mg/day p.o. and posaconazole 0.3 g/day p.o. for 4 weeks) |
| D133 | Outpatient | HCMV DNA negative, high level of CRP and persistent low level of absolute lymphocyte count relieved | Regular follow-up visits |

Abbreviations: D day, CRP C-reactive protein, SAA Serum amyloid A, IV intravenous, p.o per os, CT computed tomographic, BALF bronchoalveolar lavage fluid, mNGS metagenomics next generation sequencing, HCMV human cytomegalovirus, EBV Epstein-Barr virus, GM galactomannan

## 3 **MATERIALS AND METHODS**

The authors confirm that the ethical policies of the journal, as noted on the journal’s author guidelines page, have been adhered to and that appropriate ethical review committee approval has been received.

### 3.1 **Fungal pathogen identification**

Microbiological tests were implemented with pleural fluid, puncture drainage tubes and bronchoalveolar lavage fluid (BALF). The isolates obtained were identified by examination of micromorphological and macromorphological characteristics. After pretreatment with 70% formic acid and α-cyano-4-hydroxycinnamic acid (HCCA, Sigma‒Aldrich). The strains were identified by matrix-assisted laser desorption/ionization (MALDI) time-of-flight (TOF) mass spectrometry (MALDI-TOF MS, Zybio EXQ2000, Zhongyuan Huiji Biotechnology Co. Ltd.) according to the standard operation protocol.^12^ Mycelia were mechanically disrupted by liquid nitrogen grinding, followed by genomic DNA extraction using QIAGEN’s DNeasy® Plant Kit.^13^ The internal transcribed spacer region (ITS) was amplified with the primer pair ITS1 (5’-TCCGTAGGTGAACCTGCGG-3’) and ITS4 (5’-TCCTCCGCTTATTGATATGC-3’). DNA sequences were analysed using NCBI BLAST (https://blast.ncbi.nlm.nih.gov/Blast.cgi).

### 3.2 **Antifungal susceptibility testing**

*In vitro* susceptibility was tested according to the guidelines presented in document M38-A2 of the Clinical and Laboratory Standards Institute (CLSI).^14^ The minimum inhibitory concentration (MIC) was determined visually as the concentration that resulted in 100% inhibition. The minimal effective concentration (MEC) endpoint was taken as the lowest concentration at which the visual growth pattern change from granular to filamentous growth was detected, microscopically seen as restricted hyphal growth.^14,15^

### 3.3 **Flow cytometry analysis**

To monitor lymphocyte subsets and programmed cell death protein 1 (PD-1) expression in peripheral blood and pleural effusion to evaluate the efficacy of therapy with thymosin-α-1 and intravenous immunoglobulin, simultaneous identification and enumeration of T lymphocytes in whole blood and multicolour flow cytometric analysis of lymphocyte subsets as well as CD4+ and CD8+ T-cell subset ratios were determined using an automated AQUIOS cytometer (Beckman Coulter), as per the manufacturer’s instructions for use. Each site had an instrument and all needed reagents. Ten millilitres of pleural fluid was passed through a 40 µm filter, and the cellular pellet was spun down. Then, red blood cells (RBCs) were lysed using a soft RBC lysing solution (Beckman Coulter). Cellular viability was then assessed using fluorescent reactive dye.^16^

### 3.4 **Literature review**

We performed a literature search of *Scopulariopsis* or *Microascus* pleural infection and *Scopulariopsis gracilis* or *Microascus gracilis* infection published from 1962 to 2023 via the PubMed database (https://pubmed.ncbi.nlm.nih.gov/). In addition, the references cited in these reports have been reviewed to identify additional cases that were not found in the PubMed database. The review included an assessment of all demographic features, including year/nation, age/gender, underlying disease, transplant type, systemic antifungal agents before infection, involved sites, identified fungal organisms, treatment and therapeutic outcomes. Only those cases that met the criteria for “proven invasive fungal disease” caused by moulds as described by the consensus group of the European Organization for Research and Treatment of Cancer/Invasive Fungal Infections Cooperative Group and the National Institute of Allergy and Infectious Diseases Mycoses Study Group (EORTC/MSG) were included in this evaluation.^17^

## 4 **RESULTS**

### 4.1 *M. gracilis* and *A. fumigatus* were identified according to classical morphology, mass spectrometry and molecular biology methods

Colonies grown on SDA were first visible on Day 2 of incubation and developed into small mould colonies over the next few days (Figure 2C). The colonies were initially pale but developed a velvety and olivaceous grey colour after 2 weeks. Lacto-phenol cotton blue mounts of the colonies showed conidiophores mostly irregularly branched, usually consisting of clusters of 2 to 3 annellides borne on short branches. Annellides lageniform or somewhat cylindrical with a slightly swollen base. Conidia subspherical to ellipsoidal, with truncate base and rounded or pointed aped, arranged in long chains (Figure 2D). *M. gracilis* and *A. fumigatus* were identified by mycological characteristics, MALDI-TOF MS (Figure 2E) and ITS-based sequencing (accession no. OR131328).

### 4.2 *M. gracilis* manifests as a multidrug-resistant strain

The results revealed that multiple antifungal agents were inactive against *M. gracilis*, with MIC values of 4 μg/ml for amphotericin B (AMB), 8 μg/ml for VRC, > 8 μg/ml for POS, >16 μg/ml for itraconazole (ITC), >64 μg/ml for 5-flucytosine (5-FC), >256 μg/ml for FLZ, and minimal effective concentration (MEC) values all > 8 μg/ml for echinocandins. The susceptibility profiles of *A. fumigatus* showed POS 0.06 μg/ml, ITC 0.12 μg/ml, VRC 1 μg/ml, AmB 4 μg/ml, 5-FC >64 μg/ml and echinocandins all 8 μg/ml.

### 4.3 **Adjuvant immunomodulatory treatment successfully increased the levels of CD3+ T and CD4+ T cells while decreasing the levels of CD3+PD-1+ T and CD4+PD-1+ T cells**

The inflammatory indicators, lymphocyte subsets and PD-1 expression in peripheral blood and pleural effusion were evaluated before and after adjuvant immunomodulatory treatment with thymosin-α-1 and intravenous immunoglobulin. The levels of CD3+ T cells and CD4+ T cells were alleviated (Figure 3), while CD3+PD-1+ T cells and CD4+PD-1+ T cells were all decreased significantly both in blood (Figure 4) and pleural effusion (Figure 5) after the immunomodulatory agents were administered. Other abnormal biochemical biomarkers, such as hsCRP and IL-6 in peripheral blood and LDH, ADA, IL-1β and IL-6 in pleural diffusion, all declined substantially. (Table 2)

**FIGURE 3.**
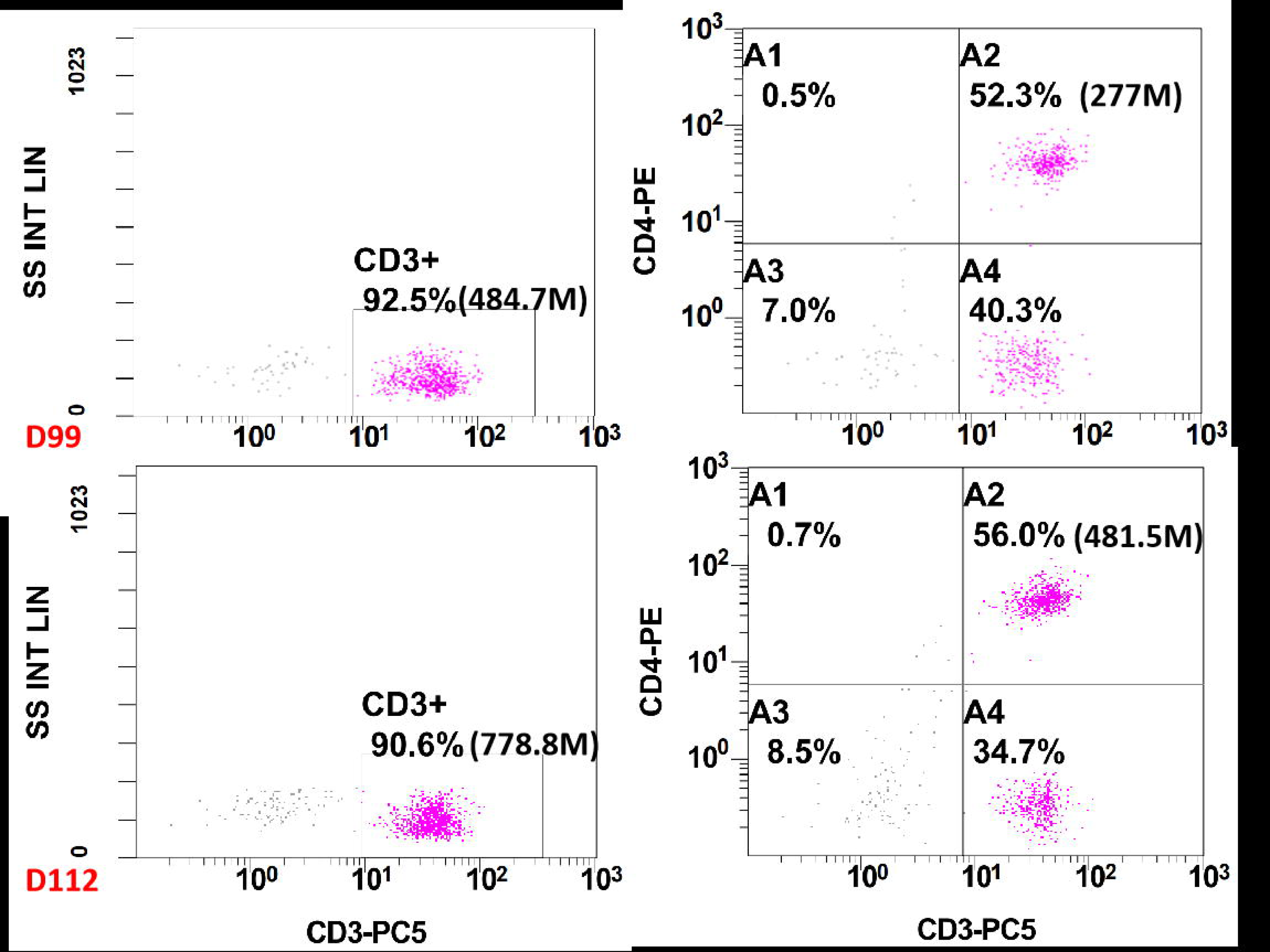
Flow cytometry results displayed the level of CD3+ T cells and CD4+ T cells were alleviated in peripheral blood after the immunomodulatory agents administered.

**FIGURE 4.**
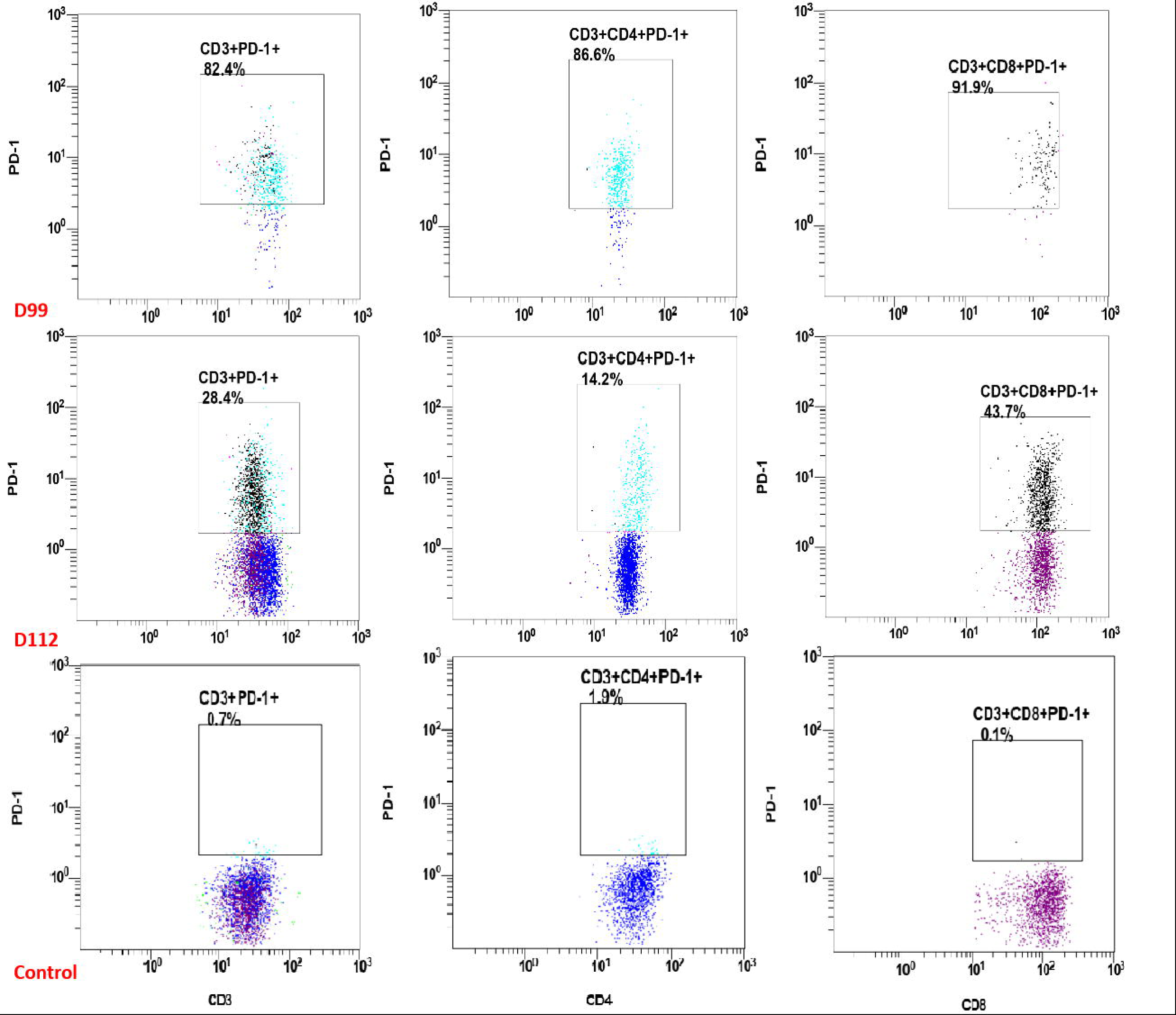
Flow cytometry results reveals the level of CD3+PD-1+ T cells and CD4+PD-1+ T cells were all decreased significantly in peripheral blood after the immunomodulatory agents administered.

**FIGURE 5.**
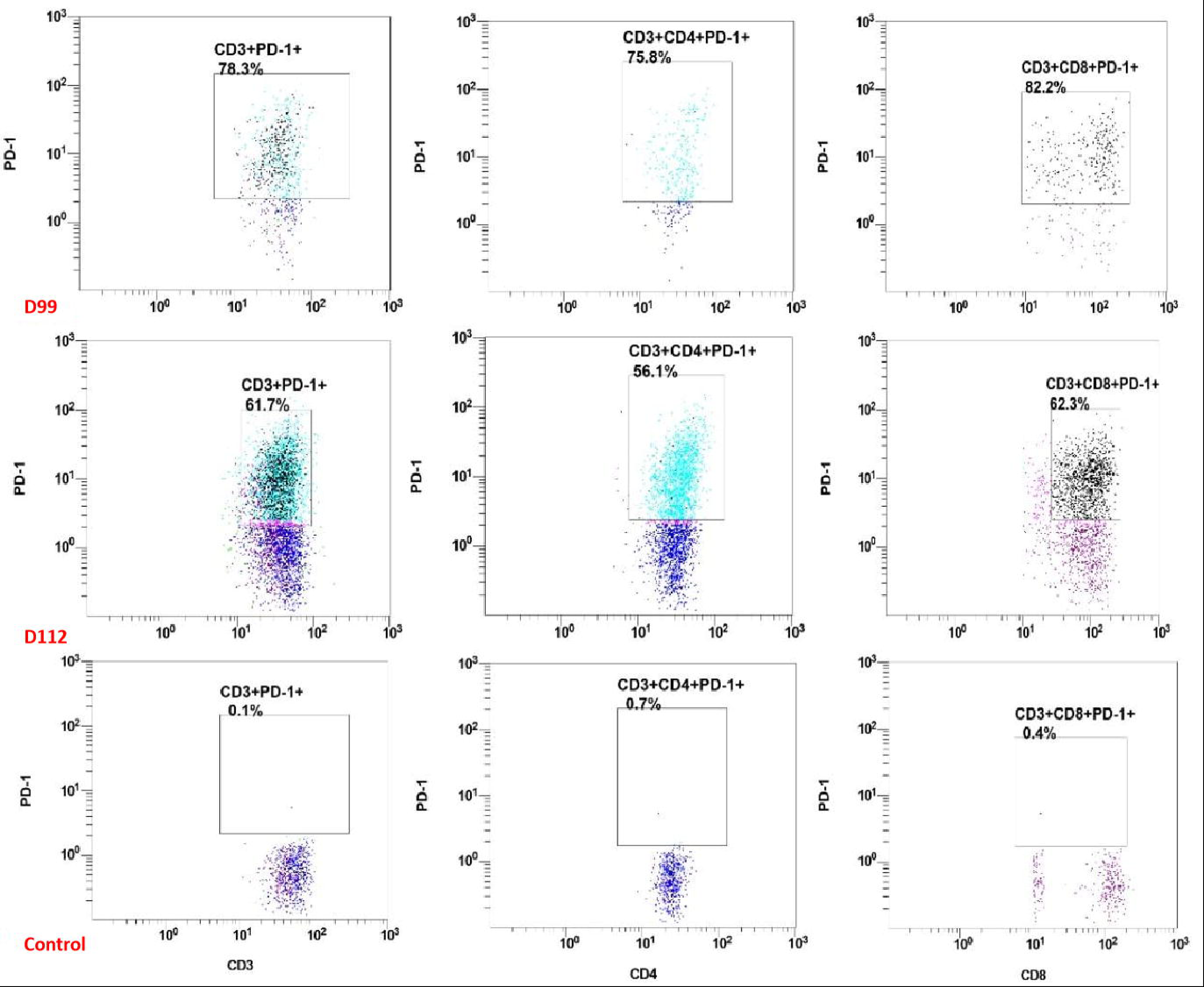
Flow cytometry results reveals the level of CD3+PD-1+ T cells and CD4+PD-1+ T cells were all decreased significantly in pleural effusion after the immunomodulatory agents administered.

**TABLE 2.** Comparison of abnormal biochemical and immunological indicators before and after the administration ofimmunomodulatory regimens.

| Time | Peripheral blood |  |  |  |  |  | Pleural effusion |  |  |  |  |  |  |  |
| --- | --- | --- | --- | --- | --- | --- | --- | --- | --- | --- | --- | --- | --- | --- |
| | CD3+T<br>($\times 10^6$/L) | CD4+T<br>($\times 10^6$/L) | CD3+PD-1+<br>(%) | CD4+PD-1+<br>(%) | hsCRP<br>(mg/L) | IL-6<br>(pg/mL) | CD3+T<br>(n) | CD4+T<br>(n) | CD3+PD-1+<br>(%) | CD4+PD-1+<br>(%) | IL-6<br>(pg/mL) | IL-1β<br>(pg/mL) | LDH<br>(U/L) | ADA<br>(U/L) |
| D99 | 484.7 | 277 | 82.4 | 86.6 | 130 | 76.91 | 737 | 355 | 78.3 | 75.8 | >5000 | 389 | 1207 | 22.4 |
| D112 | 778.8 | 481.5 | 28.4 | 14.2 | 7.0 | 36.02 | 4866 | 2818 | 61.7 | 56.1 | 510 | 73.6 | 192 | 2.5 |

### 4.4 **Literature review**

A review of the English literature since 1962 identified 5 cases reported as proven pleural *Scopulariopsis/Microascus* infections.^18–22^ The reported cases listed in Table 3 had a median age of 52.8 years (range 27-70 years) and represented 6 males. Of these cases, 2 were from the USA, and 1 each was from Belgium, India, France and China. Of these patients, 80% had solid organ transplant (SOT), and all were associated with lung transplantation. Sixty percent of patients were treated with azole antifungal prophylaxis. Because of the isolate morphologically identified as *M.cinereus* was found after sequencing to be from *M. gracilis*.^4,10^ The most commonly identified species was *M. gracilis* (3/6, 50%), followed by *S. brumptii* (2/6, 33.3%) and *S. acremonium* (1/6, 16.7%). Combined therapies were performed when diagnostic clues were found, but the therapeutic outcomes were worse, as 83.3% of patients died.

**TABLE 3.** Demographic features of reported cases with indicated pleural infection by *Scopulariopsis!Microascus* species.

| Refer<br>ence | Diagnos<br>is year/<br>nation | Age/g<br>ender | Underlying<br>disease | Transplant<br>type | Systemic<br>antifungal<br>agents before<br>infection | Involved sites | Infection<br>species | Treatment | Outcome |
| --- | --- | --- | --- | --- | --- | --- | --- | --- | --- |
| 19 | 2005/<br>Belgium | 63/M | COPD | SOT<br>(single-lung) | AMB | Heart, thyroid gland,<br>stomach, kidneys,<br>lungs | *S. acremonium* | VOR/CAS | Expired (1 day) |
| 20 | 2007/<br>India | 27/M | AIDS | —— | None | Pleural fluid | *S. brumptii* | —— | Expired cardiovascular<br>dysfunctions (14 days) |
| 21 | 2009/<br>France | 36/M | CF/RF/DM | SOT (heart<br>and lungs) | FCZ | Pleural fluid,<br>intrapericardial fluid,<br>blood clots, bronchial<br>secretions | *M. gracilis*<br>(*M. cirrosus*) | VOR/CAS | Expired <u>multiorgan</u><br>failure (9 days) |
| 22 | 2014/<br>USA | 56/M | IPF/DM | SOT (lungs) | ITC | Pulmonary pleura,<br>pleural fluid, skin,<br>ribs, sternum | *S. brumptii* | MIC/VOR/Am<br>B/TER | Expired Sepsis (48<br>days) |
| 23 | 2020/<br>USA | 65/M | IPF/PH/DM/<br>HTN/CAD/<br>HLD | SOT (lungs) | AmB/VOR | Lung, pleura, heart,<br>brain, potential eyes | *M. gracilis* | AmB/MIC/ISO/<br>TER | Expired (18 days) |
| This<br>case | 2023/<br>China | >65/M | NS/ILD/DM | —— | VOR | Pleural fluid | *M. gracilis* | CAS/POS/TEB | Cured |

Abbreviations: M, male; COPD, chronic obstructive pulmonary disease; SOT, solid organ transplant; AmB, amphotericin; VOR, voriconazole; CAS, caspofungin; AIDS, acquired immune deficiency syndrome; CF, cystic fibrosis; RF, renal failure; DM, diabetes mellitus; FCZ, fluconazole; IPF, idiopathic pulmonary fibrosis; ITC, itraconazole; MIC, micafungin; TER, terbinafine; PH, pulmonary hypertension; HTN, hypertension; CAD, coronary artery disease; HLD, hyperlipidaemia; ISO, isavuconazole; NS, nephrotic syndrome; ILD, interstitial lung disease; POS, posaconazole.

To date, only six proven cases of *M. gracilis* infections have been reported, including two cases of disseminated infection,^20,22^ two cases of invasive bronchopulmonary infection,^23,24^ and each case of primary subcutaneous infection^25^ and keratitis.^26^ A total of 83.3% of cases were underlying diseases that compromised the immune system locally or systemically. A total of 66.7% of patients also received azole antifungal prophylaxis. In addition, 66.7% were cured successfully, while the salvage treatment for two disseminated patients failed.

## 5 **DISCUSSION**

*M. gracilis*, the teleomorph of *Scopulariopsis*/*Microascus spp*., is a rare opportunistic fungus associated with human disease and is commonly found in soil, decaying organic matter and in-house environments.^4,7,8,10,22–26^ There is a paucity of information available concerning clinical infections caused by *M. gracilis*. Among the ninety-seven clinical strains morphologically identified as *Scopulariopsis*/*Microascus* spp., *M. gracilis* was the second most commonly isolated species and was most frequently isolated from BALF and sputum samples.^8^ Most of them, according to the clinical features, were determined to have fungal colonization.^8^ The diagnosis of *Scopulariopsis*/*Microascus* infections is challenging because of the delay in microbiological study completion.^4^ Precise and timely identification of these pathogens to species can be extremely important when recovered from high-risk patients. However, morphological conclusion requires sporulating culture, which considerably delays the diagnosis, while the developed DNA-based assays for identification are rapid and reliable.^27^ Typically, the isolate morphologically identified as *M.cinereus* was found after sequencing to be from *M. gracilis*.^20^ Our strain accurately identified by morphological features, mass spectrometry and ITS-based sequencing is the seventh invasive *M. gracilis* infection, as well as the sixth pleural infection caused by *Scopulariopsis*/*Microascus* spp.. Notably, in comparison to DNA-based assays, mass spectrometry analysis based on the updated database offers a cost/time-saving and accurate clinical working protocol for identifying both the genus and species levels of the *Scopulariopsis*/*Microascus* genera.

The development of spontaneous hydropneumothorax is a complication of post-COVID-19 pneumonia due to rupture of the small airway.^28^ A history of recent COVID-19 infection is a risk factor for pleuropulmonary infections, particularly in patients with comorbidities, structural defects such as bronchopleural fistula, and immunosuppressive therapies.^29^ The presence of *M. gracilis* may represent colonization or may cause localized infections at the usual residential location of the superficial tissue or the lungs.^22,30^ As the most representative aetiologic agent in *Scopulariopsis*/*Microascus spp*. infection of the pleura in extremely limitated data, *M. gracilis* occurs less frequently but causes refractory and fatal disseminated infections. Opportunistic infections of this extremely rare fungus have been reported mostly in immunocompromised patients, with most severe infections occurring in solid organ transplant patients, particularly lung transplant patients.^18,20–24^ We suspected that treatment with corticosteroids for nephrotic syndrome might have transiently suppressed immunity, resulting in opportunistic infection. Prognosis is poor, and mortality is high in immunosuppressed patients.^8,31^ Furthermore, it is worth mentioning that more than 60% of patients acquired the infection when they were under antifungal prophylaxis with azole agents. The selective pressure of antifungal prophylaxis may also contribute to the appearance of rarely invasive fungal infections, caused by moulds that are often intrinsically resistant to some classes of antifungals, which have been described as opportunistic pathogens in patients with a variety of underlying diseases.^32–34^ *Pneumocystis* pneumonia most often occurs in immunocompromised hosts, with intensified or prolonged immunosuppression, notably with corticosteroids and subsequent cytomegalovirus (CMV) infections.^35^ Coinfection with CMV may contribute to further immune dysregulation in haematopoietic cell transplant (HCT) and solid organ transplant (SOT) recipients.^36^ A total of 42.9% of SOT patients had a proven CMV infection that occurred in concurrent infection with *Scopulariopsis*/*Microascus* spp.,^18,23,37,38^ possibly because CMV can affect several components of the defence system and, therefore, could enhance the pathogenicity of other infectious agents.^18,23,39^ It is worth noting that our patient experienced a persistent low absolute lymphocyte count. Based on the analysis of lymphocyte subsets in peripheral blood and pleural effusion through flow cytometry assays, the number of T lymphocytes was significantly decreased, and T-cell exhaustion occurred. The host cannot control infections effectively because of T-cell exhaustion.^40^ PD-1 together with programmed death-ligand 1 (PD-L1) is a key biomarker of lymphocyte depletion,^41^ and upregulation of PD-1 in exhausted T cells as an immunosuppressive receptor was observed during the progression of symptomatic stages of infections, which contributes to the evolution of the severe form of the symptoms. Prognosis largely depends on the immune status of the patient, and immune reconstitution is considered essential when combating invasive fungal infections.^8^ In conclusion, elevated PD-1 levels in CD4+ T cells were associated with the inhibition of cell proliferation and decreased effector functions;^42^ in contrast, blocking or downregulating PD-1 reversed exhausted T cells, and immunosuppressive status resulted in recovery and enhancement of host innate immunity.

The potential origin of infection is vastly diverse. Outbreaks of fungal keratitis and invasive disease due to contaminated grafts used to store the issued corneoscleral button,^26^ soil contamination^43,44^ or inhaled pollution,^45^, respectively, have been described in immunocompetent hosts. In addition to the severe immunocompromised host susceptible to invasion of the opportunistic *Scopulariopsis*/*Microascus*, especially in those patients who had undergone solid organ transplants, transmission of the fungus from the donor was possible,^4,21^ and the fungus might have been accessed into the pleural space during prehospital placement of thoracostomy tubes^21^ or introduced into the skin and then into the vessels through trauma at a catheter site.^46^ Our patient was another case of catheter-related pleural infection contaminated by the indoor environment. To minimize the fungal equivalent and avoid persistent infection, the contaminated catheter should be removed immediately. The delayed diagnosis and the high rates of resistance of *Scopulariopsis/Microascus* to practically all current antifungals ^8,31,47,48^ are responsible for the high mortality associated with disseminated infections.^18,20,21,22^ A spread of pulmonary *Scopulariopsis*/*Microascus* to the pleura is unlikely in the present case because only pleuritis was detected without apparent pulmonary *Scopulariopsis*/*Microascus*.

Currently, fungal empyema therapy is not protocolized, and combinations that include available agents can be used due to the variable penetration of systemically administered antifungals into the pleural cavity.^49^ First-line treatment for systemic fungal infections, AMB and VRC, and other antifungal agents are ineffective against *Scopulariopsis*/*Microascus* spp. *in vitro*. The lack of correlation between *in vitro* and *in vivo* studies and the intrinsic resistance of fungal pathogens to many of the available antifungals limits successful therapeutic options.^50^ Susceptibility testing using CLSI or EUCAST methodologies usually shows high MIC values for all currently approved antifungal agents.^8,47^ No cut-off values have been determined, as there are insufficient data to correlate MIC values and clinical outcomes.^51^ Nevertheless, *in vitro* synergy has been demonstrated with combinations of POS and TEB, POS and caspofungin (CAS), AMB and CAS, and three drugs in combination.^30,52^ L-AMB (3–10 mg/kg/day) and VRC,^52,53^ VRC alone (i.v. and then oral step-down therapy), and lipid formulations of AMB with other antifungal agents can be moderately recommended.^52^ VRC with an echinocandin and TEB is marginally recommended.^20,52^ Limited literature on *M. gracilis* indicates higher MICs *in vitro*, ^20,22,23,25,30,48^ approximately 85.7% of the isolates had high concentrations for AmB, CAS, VOR and POS, except one case had relatively low concentrations for AMB,^22^ CAS,^48^ VOR and POS,^23^ respectively. All strains had high MIC values for FLZ, ITC, and 5-FC. The MEC values of micafungin were 0.25^23^ and 1 μg/ml,^22^ and the MIC of isavuconazole was established at 2 μg/ml.^22,54^ It is worth mentioning that the value of TEB fluctuated obviously from 1 to 16 μg/ml. The fractional inhibitory concentration indices (FICIs) range of CAS, POS, and TRB combination against *M. gracilis* was from 0.321 to 0.320, which appear to be promising, with a synergistic effect on *M. gracilis*^.30^ The MIC, POS, and TRB combination in invasive infections warranted to be an effective therapy in clinical practice.^23,24^ The other therapeutic strategies include nebulized micafungin for the treatment of *Scopulariopsis/Microascus* tracheobronchitis,^55^ combination of treatment for *Scopulariopsis/Microascus* pneumonia, which included surgery, triple combination of antifungal drugs, and granulocyte-colony stimulating factor.^56^ Moreover, an investigational antifungal within the orotomide class that is currently in late-stage clinical development, olorofim (formerly F901318), demonstrated good *in vitro* activity against the majority of *Scopulariopsis/Microascus* isolates tested, with an MIC90 value of 0.125 mg/L and a modal MIC of 0.03 mg/L.^57^ Surgical debridement (where feasible) and reversal of the host’s innate immunodeficiency are critical to survival and are strongly recommended.^52^ Particularly, for immunocompromised patients, intravenous immunoglobulin was essential to reverse the immunological crisis and was obtained with highly effective clinical outcomes.^23^ Although there is limited information about thymosin α1 (Tα1) as adjuvant immunomodulatory therapy, either used alone or combined with other treatments, the favourable combination of Tα1 with an anti-PD-1 antibody has already been postulated experimentally. Tα1 may be a treatment option for invasive fungal infection in immunocompromised hosts, but efficacy and safety data remain limited. Our immunocompetent patient was initially treated with CAS combined with POS; when the symptoms and critical biological index were relieved, the POS and TRB combination started as a sequential therapy until he was cured. Moreover, a combination of antifungal therapy with immunomodulatory therapy was shown to be an effective intervention.

In summary, the patient’s underlying comorbidities, history of recent COVID-19 infection, immunosuppressive therapy, antifungal prophylaxis with azole drugs, cytomegaloviremia, lymphocyte depletion and indwelling catheter may have played a role in the pleural seeding of *M. gracilis*. Fortunately, our patient was beneficial for successful management in a multimodal manner, including reversal or revocation of underlying predisposing factors, early administration of suitable antifungal agents, and adjuvant immunomodulatory treatment in which his condition did not progress into disseminated *Scopulariopsis*/*Microascus*. Although infection with *M. gracilis* is uncommon and rarely pathogenic in the pleural cavity, clinicians and microbiologists should be aware of the potential of *M. gracilis* infection in immunocompromised individuals at risk of opportunistic infections. Early and definitive diagnosis thorough direct microscopic testing with calcofluor white, as well as MOLDI-TOF MS, which has been suggested as a valuable tool for identification at the genus and the species level, is the key to rapidly recognize this rare pathogenic fungus. Achieving an accurate diagnosis along with appropriate antifungal management is vital for improving survival.

## AUTHOR CONTRIBUTIONS

**Zhi-Min Hu**: Investigation; methodology; formal analysis; writing–review and editing; writing – original draft; resources; visualization; data curation. **Li-Na Mao**: collected and interpreted the clinical data and wrote the manuscript. **Ti-Ying Deng**: Investigation; formal analysis; resources. **Bin-Tao Su**: revised the manuscript critically for important content. **Yi Yang**: carried out the radiological examination. **Bi-Lin Dong**: carried out the microbiological examination and nucleotide sequencing. **Qing Xu**: Investigation; formal analysis. **Shuo Yang**: Methodology; supervision; project administration; writing – review and editing. All the authors have read and approved the final manuscript.

## Data Availability

All data produced in the present work are contained in the manuscript.

## ACKNOWLEDGEMENTS

This work was supported by the Wuhan Health Scientifc Research Key Project (No.WX18A06), and the Wuhan Health Scientifc Research Program(No.WH21C21).

